# Characterisation of the retinal phenotype using multimodal imaging in novel compound heterozygote variants of *CYP2U1*

**DOI:** 10.1101/2023.03.07.23286486

**Authors:** Ferenc B Sallo, Chantal Dysli, Franz Josef Holzer, Emmanuelle Ranza, Michel Guipponi, Stylianos E Antonarakis, Francis L Munier, Alan C Bird, Daniel F Schorderet, Beatrice Rossillion, Veronika Vaclavik

## Abstract

**Purpose:** To report the retinal phenotype in two patients simulating type 2 macular telangiectasis with new variants in *CYP2U1* implicated in Hereditary Spastic Paraplegia type 56 (HSP 56).

**Methods:** Five members of a non-consanguineous family (parents and three male children) were investigated. All family members underwent a full ophthalmological evaluation and multimodal retinal imaging. Two family members demonstrating retinal anomalies underwent additional OCT angiography, dual wavelength autofluorescence and fluorescence lifetime imaging ophthalmoscopy, kinetic perimetry, fundus-correlated microperimetry, electroretinography and electro-oculography. Whole exome sequencing was performed in all five family members.

**Results:** The two siblings with compound heterozygous novel variants c.452C>T; p.(Pro151Leu), c.943C>T; p.(Gln315Ter) in *CYP2U1* demonstrated parafoveal loss of retinal transparency and hyperreflectivity to blue light, redistribution of luteal pigment to the parafoveal edge, photoreceptor loss, FLIO anomalies: a pattern compatible with that seen in macular telangiectasia type 2 (MacTel). One had manifest neurological abnormalities since early childhood, the second had no neurological abnormalities. Each parent and the third sibling were heterozygous for one variant and were neurologically and ophthalmologically normal.

**Conclusion:** These *CYP2U1* variants are associated with a retinal phenotype very similar to that otherwise specific for MacTel, suggestive of possible links in the aetiology/pathogenesis of these diseases.

## INTRODUCTION

Hereditary spastic paraplegia (HSP), also known as Strümpell-Lorrain disease, is a neurological disease characterized by either an isolated pyramidal syndrome (pure form) or in association with other neurological or extra neurological signs (complex form).^1^ Pure HSP typically presents with progressive bilateral lower leg spasticity and weakness. HSPs are among the most clinically and genetically heterogenous group of inherited neurodegenerative disorders. HSPs are transmitted by all Mendelian forms of inheritance, including de novo mutations. More than 70 causative genes and/or loci have been identified so far.^2^

Hereditary spastic paraplegia 56 (SPG56 MIM 615030, also known as SPG49), is an autosomal recessive form of HSP due to biallelic pathogenic variants in *CYP2U1* (MIM # 610670), that may manifest as a pure or as a complex form. *CYP2U1* encodes a member of the cytochrome P450 group of enzymes. This protein is involved in lipid metabolism and has also been implicated in mitochondrial function.^3^ The phenotypic spectrum of SPG56 has been expanded since the first description, to date 32 affected individuals have been reported with pathogenic variants in *CYP2U1*.^4-14^ Additional clinical features may include cerebellar ataxia, dystonia, intellectual disability, cognitive delay, visual impairment, and subclinical peripheral neuropathy. Brain Magnetic Resonance Imaging (MRI) may (rarely) show a thin corpus callosum, calcification of basal ganglia and delayed myelination.^5, 6, 12, 13^ Although visual impairment is a frequent complication of HSPs,^15^ a pigmentary maculopathy has only been described in a few families^5, 10^ and isolated cases.^7, 16^

Macular telangiectasia type 2 (MacTel) is a progressive degenerative neuroglial-vascular macular disease that leads to loss of central vision, with a late onset of symptoms and a characteristic constellation of retinal signs.^17-21^ The condition has a strong genetic component, supported by extended families with multiple affected individuals, linkage, and genome-wide association studies, however, the inheritance pattern is not clear because of the variable penetrance and expressivity of the disease.^22-25^

In this study we aim to characterize the retinal phenotype of two male siblings with two novel pathogenic and likely pathogenic *CYP2U1* variants, to investigate the similarities of their retinal phenotype to that of MacTel and to discuss the potential implications.

## PATIENTS AND METHODS

The protocol of the study adhered to the tenets of the Declaration of Helsinki and was approved by CER-VD, (Canton Ethics committee for research on human, Lausanne, Switzerland) Req: 2019-00518. Written informed consent was obtained from all patients prior to inclusion in the study, as well as their consent for publication of the results.

### Patient selection and clinical evaluation

Five members of a Swiss, non-consanguineous family were examined (Figure 1). At the time of baseline ophthalmic examinations, the father (I.1) was and mother (I.2) were in their mid 50s, the eldest sibling (II.1) in mid twenties, siblings II.2 and II.3 in their early 20s, all siblings were male. The eldest sibling (II.1) developed gait abnormalities at an early age and was diagnosed with Strümpell-Lorraine disease, prompting a genetic screening of the whole family.

**Figure 1.**
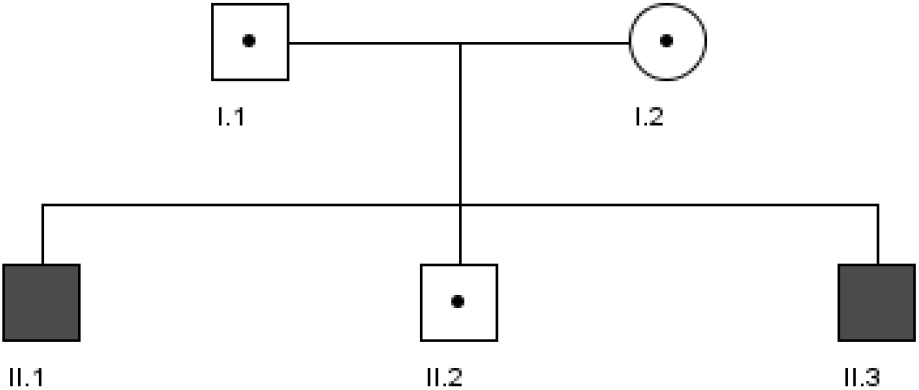
Pedigree of the family. Filled symbols represent siblings affected by maculopathy. Dots represent CYP2U1 carrier states.

### Whole Exome Sequencing

Whole exome sequence of Individual II.1 was performed using the following procedure: DNA was captured, and coding regions and splice sites were enriched using the Agilent SureSelect QXT Human All Exon V5 capture kit. Sequencing was performed on an Illumina instrument following the manufacturer’s protocol. Targeted bioinformatics analysis of a panel of 137 genes involved in spastic paraplegia (HSP_v1.00_137 genes_ER.txt_.HUGO) was done through locally developed pipelines, integrating BWA v0.7.10, Picard 1.80, GATK 3.50, ANNOVAR vSept2015. We selected only the variants from the genes of interest, masking the rest of the data. The evaluation of variants used the following databases: dbSNPv142, ExAC 0.3, gnomAD 2.0, ClinVar 2016, HGMD 2016, LOVD, and the local database of variants. Prediction programs used were: SIFT, Polyphen2, MutationTaster and dbscsnv11 and HSF v.3.0. Variants were classified according to the recommendations of the American College of Medical Genetics.^26^ Variants detected were validated and familial segregation was performed by Sanger sequencing. The same variants were validated in patient II.3.

### Ophthalmic phenotyping

All family members underwent a full ophthalmological evaluation including Goldmann applanation tonometry, slit lamp examination, indirect ophthalmoscopy and color fundus (CF) imaging, using a Topcon TRC50DX Retinal Camera (Topcon Co., Tokyo, Japan) and a Zeiss Clarus 500 Retinal Camera (Carl Zeiss Meditec AG, Jena, Germany). Optical Coherence Tomography (OCT), 488nm fundus autofluorescence (AF), blue light reflectance (BLR) imaging and infrared (IR) imaging were performed using Heidelberg Spectralis OCT+HRA devices (Heidelberg Engineering, Heidelberg, Germany).

Two family members with detectable retinal anomalies underwent additional retinal imaging including Optical Coherence Tomographic Angiography (OCT-A) using an AngioVue device (Optovue Inc., Fremont, California, USA). Dual Wavelength Autofluorescence (DWAF) imaging for measurement of macular pigment optical density with excitation wavelengths of 488nm (blue) and 518 (green) was performed. Additionally, Fluorescence Lifetime Imaging Ophthalmoscopy (FLIO, 473nm excitation wavelength, 498-560nm detection range) was done using custom Heidelberg Spectralis devices (Heidelberg Engineering, Heidelberg, Germany). FLIO is used to detect the decay time of natural retinal fluorophores after excitation by a blue laser light.^27, 28^ The combination of autofluorescence intensity and fluorescence lifetime distribution provides additional information, especially from weak retinal fluorophores.

### Functional testing

Functional testing included best corrected visual acuity (BCVA) measurements to Early Treatment Diabetic Retinopathy Study (ETDRS) standard at 4 meters, kinetic perimetry (Octopus 900, Haag-Streit AG Diagnostics, Koeniz, Switzerland) and automated, fundus-correlated microperimetry using a Centervue MAIA device (Icare Finland Oy, Vantaa, Finland) and a 95-point test pattern developed for the MacTel CNTF phase II study.^29^ *‘‘En face’’* OCT images of the ellipsoid zone (EZ) and microperimetric retinal sensitivity threshold data were superimposed and adjusted to attain exact correspondence. Sensitivity thresholds measured within EZ break areas were compared to those measured outside the lesions.^30^ Results are reported in decibels. Fixation stability was expressed as the bivariate contour ellipse area (BCEA), which is the area of an ellipse on the retinal surface within which the center of the target was imaged 68% of the time. BCEA is a standardized measure that provides a means for quantification and comparison of fixation stability. Smaller BCEA values correspond to more precise fixation.^31^

Full-field electroretinograms (ERG) were performed following pupil dilation (using 1% tropicamide and 2.5% phenylephrine hydrochloride) according to International Society for Clinical Electrophysiology of Vision (ISCEV) standards using an Espion Visual Electrophysiology System (Diagnosys Vision Ltd, Dublin, Ireland). The protocol included rod-specific and standard bright flash ERG, both recorded after a minimum of 20 minutes dark adaptation. Photopic 30Hz flicker cone and transient cone ERGs were recorded following 10 minutes of light adaptation.

## RESULTS

### Clinical history

Patient II.1 started autonomous walking at normal age but with abnormal plantar flexion of foot (equine’s foot, walking on his toes). He had medical follow up since then for gait abnormalities. Neurological examinations described a spastic gait with hyperextension of both knees, swaying hips and plantar flexion of the feet. He was diagnosed with Strümpell-Lorrain disease (Hereditary Spastic Paraplegia 56) in during his first years of life. To improve motor function, several surgical muscle extensions and botulinum toxin injections were performed. At baseline (mid 20s), the patient can walk autonomously, but frequently uses a wheelchair. Cerebral and spinal MRI performed at regular intervals during adolescence was within normal limits. Patient II.1 smokes cannabis on a regular basis for the relief of spasticity-related pain. He reported no subjective visual symptoms at any point in time. Patients I.1-2 (parents) and siblings II.2 and II.3 were neurologically asymptomatic. Patient II.3 had reduced visual acuity since he was a teen. He also reports photophobia starting in his 20s.

### CYP2U1 variants

Whole Exome sequencing with bioinformatics analysis of a panel of HSP genes revealed NM_183075.2: c.452C>T; p.(Pro151Leu), classified as likely pathogenic (according to ACMG criteria), and NM_183075.2: c.943C>T; p.(Gln315Ter), classified as pathogenic according to ACMG criteria. Both variants were validated by Sanger sequencing; Patients II.1 and II.3 were compound heterozygotes for both variants. Familial segregation confirmed that each parent (I.1 and I.2) is a heterozygous carrier of each variant. Patient II.2 is a carrier of the c.452C>T; p.(Pro151Leu) variant.

### Ophthalmic examinations

Slit lamp biomicroscopy revealed normal anterior segments, intraocular pressures were within normal range and optic nerve head morphology was normal in all patients examined. **Patients I.1, I.2** and **II.2** were subjectively asymptomatic, performed functionally within normal parameters and retinal imaging demonstrated no abnormalities in their phenotype.

### Patient II.1

#### Functional testing

BCVA at the time of imaging was 0.12 logMAR with the right eye and 0.14 logMAR with the left (Snellen equivalent of 0.8 with either eye). Electrophysiological testing was attempted but was not feasible due to photophobia. Goldmann kinetic visual field testing revealed normal visual fields in both eyes. MAIA microperimetry showed a mean retinal sensitivity of 25.6 dB (median 25.0 dB, SD 2.3 dB) with the right eye and 24.0 dB (median 25.0 dB, SD 3.3 dB) with the left eye. EZ breaks were too small for calculating mean sensitivity over retinal areas within the EZ lesions. Fixation stability (BCEA) was 2.2°^2^ with the right eye and 6.5°^2^ with the left eye.

#### Multimodal Retinal Imaging (Figure 2.)

CF imaging revealed a loss of retinal transparency and BLR imaging a hyperreflectivity within the parafovea in both eyes. CF and BLR imaging modalities were confounded by patchy retinal surface reflectivity. OCT imaging showed low reflective spaces in the inner retina within the parafovea, just internal to the outer nuclear layer (ONL) and internal to the inner nuclear layer (INL). Small patches of low reflective spaces were also present in the outer retina, at the edge of the parafovea, temporal and inferior to the foveal centre in the right eye, nasal to the fovea in the left eye. Blue (488nm) AF imaging showed a loss of the central peak of luteal pigment at the fovea; DWAF imaging revealed a depletion of MP within the parafovea with a ring of increased MP density at approximately 5.5 degrees radial distance from the foveal center. FLIO imaging showed short fluorescence lifetimes in the foveal center (central 1mm ETDRS subfield) with values of 250-264ps (age-corrected reference value: 135±21ps). In the inner (3mm) ETDRS-ring, the values were prolonged to 278-284ps (reference: 217±23ps), whereas in the outer ETDRS ring (6mm) and the peripheral retina the values were normal with 258-260ps (reference: 248±16ps). The ring of prolonged fluorescence lifetimes was not clearly evident in the autofluorescence intensity image and showed only a vague coincidence to the central slightly decreased autofluorescence intensity. However, it corresponds to a central area of changes in the infrared image and to a zone of irregular or missing photoreceptors in the OCT image. OCT-A imaging revealed slightly enlarged intercapillary spaces around the foveal avascular zone (FAZ) in both eyes. FAZ size was within normal parameters.

**Figure 2.**
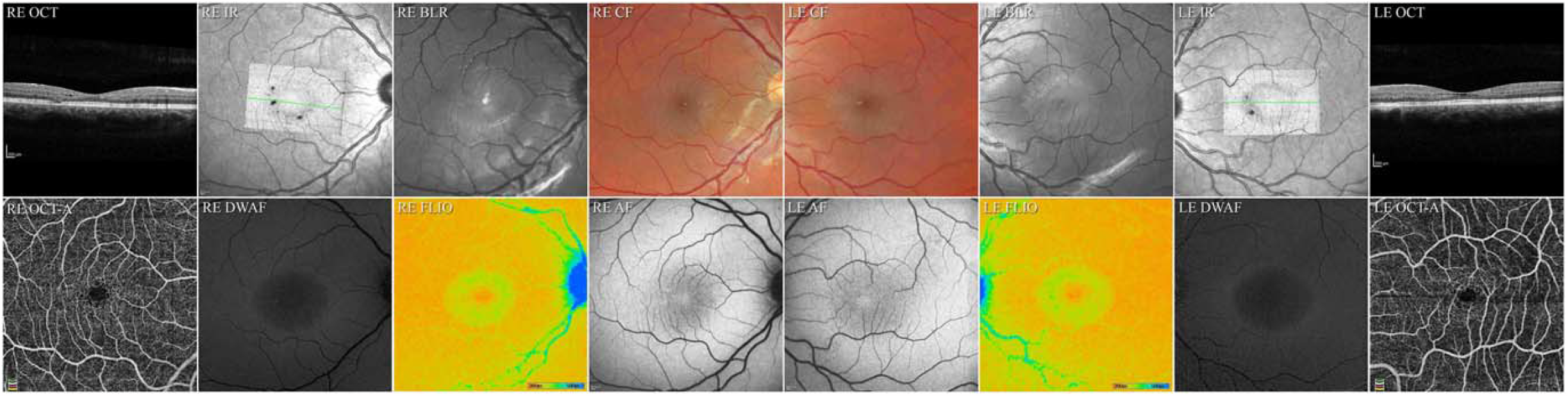
Multimodal retinal imaging of patient II.1 RE: indicates the right eye, LE the left eye, **Colour fundus** (CF) images demonstrate a loss of retinal transparency in both eyes. **Blue light reflectance** (BLR) images show an increased scatter within the same area as the loss of retinal transparency. (In the RE the bright round hyperreflectivity at the fovea and the faint round hyperreflectivity temporal inferior to the fovea are artifacts due to internal reflections within the optics of the recording device, the arcuate hyperreflectivity along the arcades in both eyes are retinal surface reflections). **Infrared** (IR) images with an overlay of the ‘*en face*’ image of the EZ and a green line marking the position of the corresponding sample **OCT B-scan** traversing the foveal center. En face images of the EZ show in both eyes an attenuation of the signal within the parafovea with an outline coinciding to those seen in BLR, DWAF and FLIO images. In the right eye multiple focal discontinuities (breaks) in the EZ are apparent within this area, temporally and inferiorly. In the left eye the EZ breaks are located nasal to the fovea, within the papillomacular area. Within the OCT B-scan of the RE, a focal discontinuity in the photoreceptor outer segment line and the ellipsoide zone are apparent with a thinning/disorganisation of the ONL as well as low reflective spaces within the inner retina. The OCT of the LE shows low reflective spaces within the inner retina and a slight attenuation of the in the photoreceptor outer segment line. The hyperreflectivity within the ONL in the nasal retinal is most likely due to an improved visibility of Henle’s layer due to a slight tilt of the scan. The **OCT angiographic** (OCT-A) images show in the right eye a slightly enlarged, in the left eye a normal size FAZ with bordering dilated capillaries and increased intercapillary gaps. **Fundus autofluorescence** (AF) images show a loss of the normally present central hypo-AF peak and increased AF on the temporal side in both eyes whereas **dual wavelength autofluorescence** (DWAF) imaging demonstrates a generalised low luteal pigment (LP) content within a ring of increased LP at approximately 6 degrees eccentricity. Fluorescence liftetime imaging (FLIO, color range: 200ps in red - to 500ps in blue) imaging shows a small central area with short fluorescence lifetimes surrounded by a clearly demarcated ring of prolonged lifetimes. The peripheral retina features a normal fluorescence lifetime distribution.

### Patient II.3

#### Functional testing

BCVA at the time of imaging was logMAR 0.1 with each eye (Snellen equivalent 0.8). ISCEV standard full-field ERGs, EOG and Goldmann kinetic visual fields were within normal limits. Using Maia microperimetry, EZ break areas showed a mean retinal sensitivity of 15.3 dB (median 17.0 dB, SD 6.9 dB) in the right eye and 16.2 dB (median 16.0 dB, SD 6.9 dB) in the left eye. Mean retinal sensitivity outside the EZ breaks was 25.7 dB (median dB, SD 3.6 dB) with the right eye and 25.9 dB (median 27.0 dB, SD 3.2 dB) with the left eye. Fixation stability (BCEA) was 0.1°^2^ with the right eye and 0.2°^2^ with the left eye.

#### Multimodal Retinal Imaging (Figure 3.)

CF imaging revealed a bilateral loss of retinal transparency within the parafovea. In the left eye, three small foci of perivascular pigmentation in the deeper layers of the retina were apparent. BLR imaging showed a hyperreflectivity within the same region as the loss of retinal transparency. CF and BLR imaging modalities were confounded by patchy retinal surface reflectivity. OCT imaging showed within the parafovea an extensive outer retinal (photoreceptor) degeneration with a loss of the photoreceptor outer segment tip (POST) and EZ lines, and thinning or loss of the ONL, topographically in a petaloid pattern centered on the fovea, with foveal sparing in both eyes. The temporal part of the parafovea appeared less affected in both eyes. Within the area of outer retinal atrophy, a disorganisation and granular hyperreflectivity was apparent within the inner retinal layers. Low reflective intraretinal spaces were apparent just external to the retinal nerve fiber layer (RNFL) and to the outer plexiform layer (OPL). Corresponding to pigment seen in CF images and to hypoAF, patches of increased scatter (hyperreflectivity) was detectable in the mid-outer retina, extending towards the retinal pigment epithelium (RPE). AF imaging showed an irregular depletion of central luteal pigment in both eyes and small hypoAF foci associated with minor blood vessels, two in the right eye, four in the left eye. In the left eye one hypoAF focus appeared to join two smaller vessels (Figure 2, LE AF488nm). DWAF imaging revealed a depletion of MP within the parafovea with a ring of increased MP density at approximately 6° radial distance from the foveal center. FLIO imaging showed short fluorescence lifetimes in the foveal center (1mm) with values of 250-260ps (age corrected reference value: 135±21ps). In the inner ETDRS-ring (3mm), the values were prolonged to 281-288ps (reference: 217±23ps), whereas in the outer ETDRS ring (6mm) and the peripheral retina the values were normal with 249-253ps (reference: 248±16ps). The outer border of the ring of prolonged fluorescence lifetimes corresponded approximately to a central area of hypoautofluorescence in the autofluorescence intensity image and correlates to the changes seen in the infrared image. However, in the AF image the hypoAF was poorly demarcated whereas in the FLIO image the outer edge of the ring was well defined. It corresponded to a zone of irregular or missing photoreceptors in the OCT image and featured a speckled pattern of irregularly prolonged lifetime areas. Outside this ring of prolonged lifetimes, an attenuated band of short lifetimes was observed. In OCT-A images, an irregular, enlarged FAZ was apparent with surrounding wider intercapillary spaces in both eyes. In the left eye, capillaries demonstrated a radial pattern converging on a focal pigmented lesion temporal inferior to the foveal centre. Historical OptoView OCT & OCT-A images were available for patient II.3, from three years prior to baseline imaging *(Figure 4.)*. EZ break area in ‘*en face’* images increased over this timespan by approximately 95% in the right eye, and 77% in the left eye. The pattern appeared radial/petaloid with a sparing of the fovea and a relative sparing of the temporal part of the parafovea. Historical IR images and B-scans from 2013 demonstrate that the EZ breaks as seen in Figure 4 A and B were already present, although the scan parameters did not permit break area measurements with reasonable accuracy.

**Figure 3.**
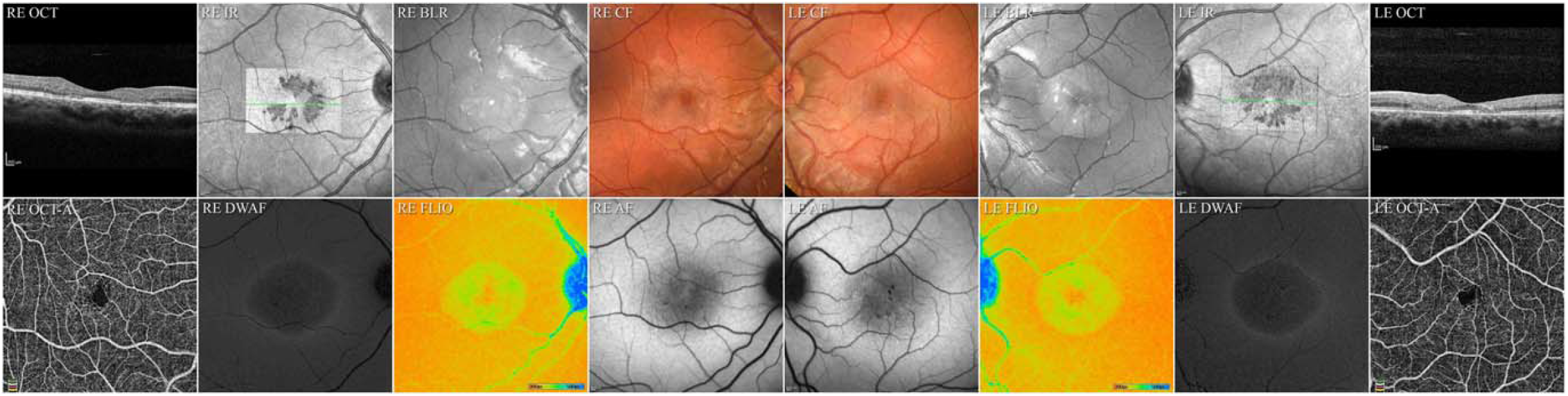
Multimodal retinal imaging of Patient II.3 RE indicates the right eye, LE the left eye, **Colour fundus** (CF) images demonstrate a loss of retinal transparency in both eyes and small brown vessel-adjacent pigment foci in the left eye. **Blue light reflectance** (BLR) images show an increased scatter within the same area as the loss of retinal transparency. (in the RE the small bright round hyperreflectivity and larger circular hyperreflectivity at the fovea are artifacts due to internal reflections within the optics of the recording device, the hyperreflectivity along the vessel arcades in both eyes are retinal surface reflections). **Infrared** (IR) images with an overlay of the ‘*en face*’ image of the EZ and a green line marking the position of the corresponding sample **OCT B-scan** traversing the foveal center. En face images of the EZ show in both eyes a radial or petaloid arrangement of breaks in the EZ and a thinning of the ONL. Notable is the relative sparing on the temporal side, especially in the right eye. Small low reflective spaces are apparent within the inner retina. The **OCT angiographic** (OCT-A) images show in both eyes an enlarged, deformed FAZ with bordering dilated capillaries and increased intercapillary gaps. **Fundus autofluorescence** (AF) images show a loss of the normally present central hypo-AF peak in both eyes. Small focal hypo-AF spots are apparent in both eyes some of these in the LE colocate with the brown focal vessel-adjacent pigment apparent in CF images. **Dual wavelength autofluorescence** (DWAF) imaging demonstrates a generalised low luteal pigment (LP) content within a ring of increased LP at approximately 7 degrees eccentricity. Fluorescence liftetime imaging (FLIO, color range: 200ps in red - to 500ps in blue) shows a slight central area with short fluorescence lifetimes surrounded by a clearly demarcated ring of prolonged lifetimes. This corresponds to the area of photoreceptor atrophy in the OCT image. The ring of prolonged fluorescence lifetimes features an external border with a small ring of short fluorescence lifetimes, fading out towards the retinal periphery, which features a normal fluorescence lifetime distribution.

**Figure 4.**
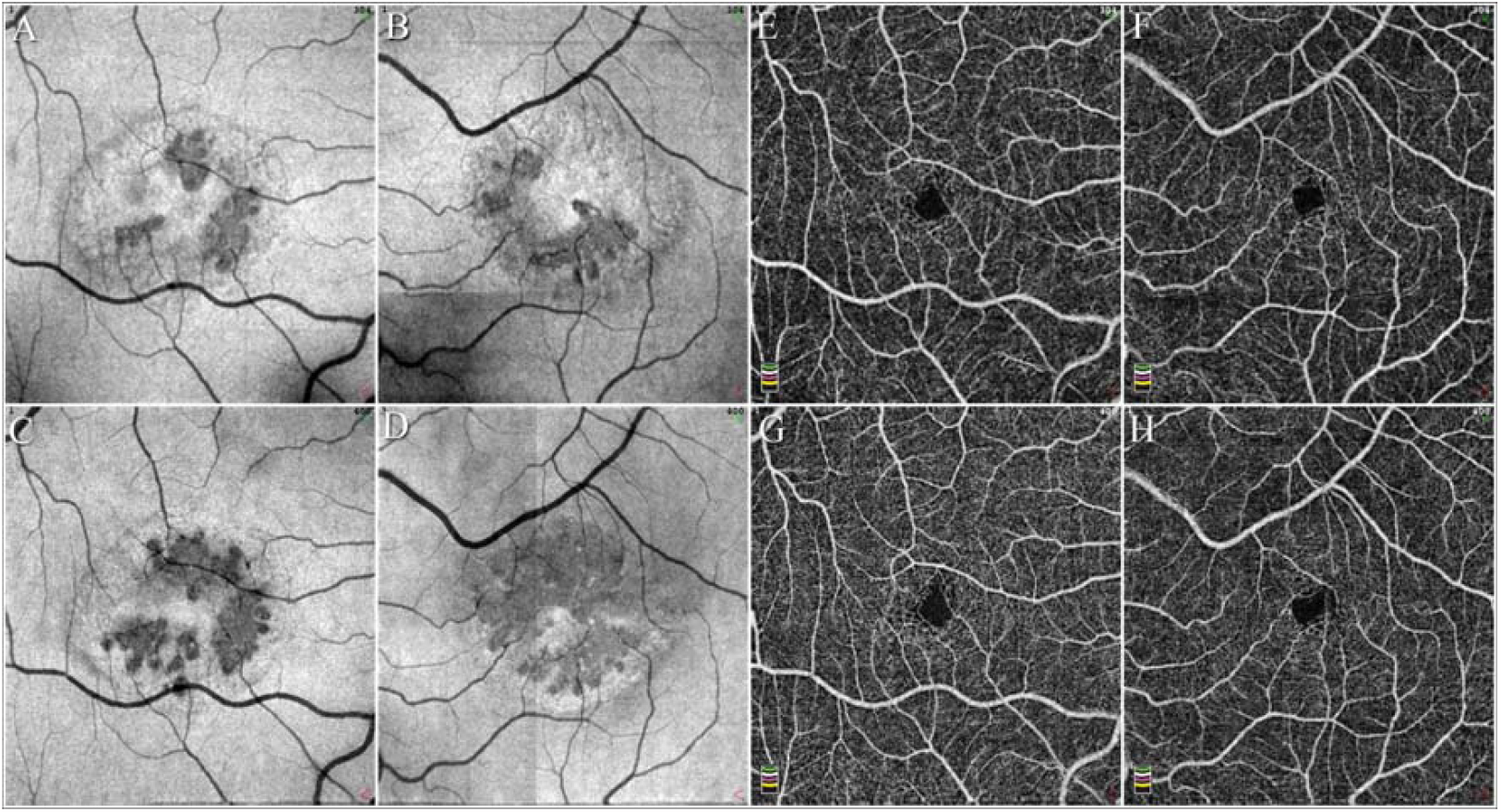
Progression patterns of neurodegenerative and vascular changes in patient II.3 A-D, *en face* OCT images of the EZ, A&C right eye, B&D left eye, E-H: OCT-A images, E&G right eye, F&H left eye, top row (A, B, E, F) three years prior to baseline, bottom row (C, D, G, H) at baseline. The EZ break apparent three years prior to baseline imaging show a radial distribution with no apparent predominance of the temporal area as in typical MacTel. At baseline the focal EZ loss appears to have progressed locally by an enlargement of the previously present foci as well as by the appearance of new foci, notably superior to the fovea in the left eye. OCT-A images show an enlargement of the FAZ over time, with an increase in intercapillary gaps and in the left eye a radial arrangement of vessels around a focal brown pigment apparent in the CF image and as a focal hypo-AF in the AF image. This feature is chararcteristic of MacTel.

## DISCUSSION

Visual impairment is a frequent complication of HSPs,^15^ optic atrophy being the most frequently described and it can also be an isolated finding.^32^ Early cataract, retinitis pigmentosa and nystagmus have also been reported.^33^ An isolated maculopathy has been described in a few cases.^5^ Leonardi et al.^10^ reported bilateral macular atrophy with RPE changes, a mild temporal paleness of the optic nerve and severely reduced BCVA in three patients with a homozygous c.1168C>T (p.R390*) variant in *SPG56/CYP2U1*. Retinal anomalies were noted before the development of motor defects. Iodice et al^7^ reported haemorrhagic macular lesions preceding a ‘pigmentary maculopathy’ in a 34 year old female with c.1288?1G>A; and c.1545_1546delTTAC in CYP2U1 however the phenotype was not described. Legrand et al.^14^ observed in three patients with homozygote variants in *CYP2U1* a maculopathy, reduced central retinal thickness and outer retinal atrophy, RPE atrophy (one patient) and a haemorrhagic CNV (one patient). The latter patient had the same homozygote *CYP2U1* c.1168C>T (p.R390*) variant as the family described by Leonardi.^10^

Our fundoscopic and OCT findings are compatible with previous reports, our cases may represent a milder form and/or an earlier stage of the retinal phenotype. The constellation of retinal anomalies of both affected patients however show striking similarities to the phenotype of MacTel.^17^

The MacTel phenotype includes characteristic neurodegenerative and vascular signs. A parafoveal loss of retinal transparency and a corresponding hyperreflective BLR pattern are early signs of MacTel.^34^ Both Patients II.1 and II.3. demonstrated these signs, Patient II.1 with the same parameters as in MacTel, whereas in II.3 the abnormal area was 30% larger.^35^ MacTel is characterized by a specific progressive pattern of MP redistribution, starting as a wedge-shaped depletion in the temporal parafovea, with a subsequent loss of the central peak and progressing toward a generalized depletion of MP within a ring of increased MP density at the edge of the parafovea. With the exception of Sjögren-Larsson syndrome,^36^ this pattern appears to be specific to MacTel. Both patients II.1 and II.3 demonstrated the late stage of this pattern. As in MacTel, the abnormal areas seen in DWAF and BLR images showed a close correspondence.^34^ FLIO values at the fovea showed a depletion rather than an absence of short lifetime fluorphores which could indicate residual MP and/or remaining central photoreceptors contributing to relatively short lifetimes.^37^ FLIO imaging showed a radially symmetrical concentric ring of prolonged lifetime values, without the temporal predominance seen in MacTel.^38^ FLIO changes colocated with OCT anomalies, but were better visible, also in some areas with apparently normal photoreceptors. Prolonged lifetimes may correlate to a long-standing degeneration of outer retinal layers.^39^ Interestingly, there was an increased density of shorter fluorescence lifetimes at the outer border of the ring, which is normally not the case at that eccentricity. As this ring corresponds to a bright area in DWAF, no MP dislocation is assumed. The origin of this remains to be investigated.

OCT signs of MacTel include hyporeflective retinal spaces and a break in the EZ. In Patient II.1, bilateral hyporeflective spaces were present in the inner retina around the foveal centre, just internal to the ONL, internal to the INL and in the outer retina, near the outer boundary of the parafovea, temporal and inferior in the right eye, however nasal to the fovea in the left eye. In MacTel all abnormal lesions tend to appear first temporal to the foveal centre, thus this pattern is different from that seen in MacTel. In patient II.3, some low reflective spaces were apparent just external to the RNFL and to the OPL, which is compatible with MacTel. The OCT phenotype dominated by an extensive loss of the outer retina is also characteristic of the later stages of MacTel. Viewed ‘en face’, the pattern of EZ loss was restricted to the parafovea (as in MacTel), the distribution however was predominantly on the nasal side, with a radial distribution and a sparing of the foveal centre and the temporal part of the parafovea. This pattern was also seen in historical images recorded three years prior to baseline. The earlier ‘seeding’ foci appeared around the fovea with some nasal predominance, progression appeared local, in a radial pattern. This is different from that seen in MacTel, which typically starts temporal to the foveal centre progressing locally in all directions.^35^

Retinal sensitivity loss over EZ break areas was similar to that seen in MacTel although of lower amplitude, i.e. the scotomata were less ‘deep’ than in MacTel.

The later stages of MacTel are characterized by perivascular brown pigment plaques in the mid-retina. In the left eye of patient II.3 small pigment foci were detectable surrounding small vessels, similar to early-mid stage MacTel.^40^

MacTel is also associated with characteristic vascular signs. Fluorescein angiography was not performed in the current study; however, OCT-A was performed, permitting an analysis of approximate vessel lumen shapes. In patient II.3, a progressive enlargement and deformation of the FAZ was notable, with increasing intercapillary spaces around the FAZ, in the left eye the vessel configuration indicated a focal lateral contraction of the retina,^41^ as frequently seen in MacTel. In patient II.1, the right eye demonstrated similar vascular changes, whereas the left eye appeared normal. Other vascular anomalies common in MacTel, like telangiectatic vessels, blunted and/or right-angle veins were not detected. Overall, neurodegenerative signs appeared to dominate the phenotype of both Patients II.1 and II.3, more than what is typical for MacTel, although similar cases have also been reported in MacTel.^42^ Secondary neovascularisation, an optional late complication of MacTel - also reported in patients with CYP2U1 mutations^7, 14^ -, was not seen in our patients.

The etiology and pathogenesis of MacTel are not fully understood. MacTel has a genetic component evidenced by extended families having multiple affected family members, however, determining a disease mechanism has been difficult due to the highly heterogenous genetic architecture, and variable penetrance and expressivity of the disease.^22, 23, 24, 43^ Recent studies have linked MacTel to low circulating levels of serine and glycine which drive a concomitant increase in a neurotoxic lipid species, deoxysphingolipids (deoxySL).^25,44, 45^ The identification of a deoxySL-linked MacTel mechanism was facilitated by the discovery that rare mutations in the serine palmitoyltransferase (SPT) genes SPTLC1 and SPTLC2 that directly elevate levels deoxySL and are causative for a rare peripheral neuropathy, hereditary sensory and autonomic neuropathy type 1 (HSAN1)^46, 47^ are also causative for MacTel. ^44, 48^ A subsequent whole exome sequencing (WES) screen identified functional variants in the gene encoding the rate-limiting serine biosynthetic enzyme, phosphoglycerate dehydrogenase (PHGDH) which causes a reduction of serine synthesis and elevation of deoxySL synthesis.^43^ These variants are rare in MacTel patients, collectively accounting for less than 4% of the disease load, however their identification was essential to uncovering a central disease mechanism linking MacTel to decreased serine to elevated deoxySL levels. It is apparent that serine levels and deoxySLs do not explain the entire functional disease mechanism and the identification of additional causative mutations is critical to understanding contributing factors to the disease. Despite extensive WES studies, causal genes and variants leading to MacTel disease have remained elusive. Like the discovery of SPT mutations, here, we present findings that identify HSP and MacTel comorbidity in patients with CYP2U1 mutations, indicating potentially new disease mechanisms.

The *CYP2U1* gene encodes a member of the cytochrome P450 family, enzymes known to play key roles in tissue-specific conversion of natural substrates into locally active hormones, vitamins and signaling molecules. Pujol et al.^4^ found in both humans and rodents that a loss of *CYP2U1* impacts mitochondrial activity essential for various processes, such as lipid import or apoptosis, whose dysfunction is closely associated with diseases.^49^ They hypothesized that SPG56 has two components: a neurodevelopmental component related to folate metabolism and a neurodegenerative component related to mitochondrial alterations with age. The CoQ alteration in SPG56 patients, presented a 12-fold increase of the oxidized form compared with controls. They also observed this increase in Cyp2u1−/− mice, and it worsened over time. CoQ constitutes a key component of the mitochondrial respiratory chain but, it also exerts an essential antioxidant role. Accumulation of the oxidized form in the absence of *CYP2U1* could enhance oxidative stress, leading to neurodegeneration and macular diseases.

The potential role of *CYP2U1* in mitochondrial function and the onset of MacTel is consistent with recent findings by Birtel et al. who reported two cases of chronic progressive external ophthalmoplegia associated with sporadic single mtDNA deletions (m.11037_14597del), demonstrating some phenotypic characteristics associated with MacTel.^50^ The authors hypothesised that MacTel may involve mitochondrial dysfunction^51^ and noted that the susceptibility loci identified by Scerri et al.^25^ include genes encoding mitochondrial enzymes, and that low serine levels and increased levels of deoxysphingolipids, have previously been linked to disrupted mitochondrial function.^44, 52, 53^

In conclusion, we demonstrate that *CYP2U1* variants are associated with a retinal phenotype very similar to that otherwise specific to MacTel, suggestive of possible links in the aetiology and/or pathogenesis of these diseases. Retinal screening of patients suffering from neuromuscular diseases may provide clues to an early diagnosis and the pathogenetic pathways involved. *CYP2U1* should be included in panels of genes involved in macular dystrophies.

## Data Availability

All data produced in the present work are contained in the manuscript

## ACKNOWLEDGEMENTS

Acknowledgments: we would like to thank the Family for their dedication and help.

